# Statistical significance misuse in public health research: an investigation of the current situation and possible solutions

**DOI:** 10.1101/2023.09.04.23295032

**Authors:** Alessandro Rovetta

## Abstract

**Background:** Despite the efforts of leading statistical authorities and experts worldwide and the inherent dangers of interpretative errors in clinical research, misuses of statistical significance remain a common practice in the field of public health. Currently, there is a need to attempt to quantify this phenomenon.

**Methods:** 97 studies were randomly selected within the PubMed database. An evaluation scale for the interpretation and presentation of statistical results (SRPS) was adopted. The maximum achievable score was 4 points. The abstracts and the full texts of the manuscripts were evaluated separately to highlight any differences in presentation. In this regard, a paired Student t-test was implemented.

**Results:** All studies failed to adopt statistical significance as a continuous measure of compatibility between the model and the test result. The vast majority of them did not provide information on the validation of the model used. However, in most cases, all results were reported in full within the manuscripts. Substantial differences (a-f) between abstracts (a) and full texts (f) were highlighted when applying the SRPS (null hypothesis, P<.0001, a-f=1.2, Cohen’s D=1.7; a-f=0.5 hypothesis, P<.0001, a-f=0.7, Cohen’s D=1.0).

**Conclusion:** When contextualized in the current scenario, these findings provide evidence of widespread and severe shortcomings in the use and interpretation of statistical significance measures in clinical and public health research during 2023. Therefore, it is essential for academic journals to compulsorily demand higher scientific quality standards.

## Introduction

### Background

Despite decades of intense informative efforts, the misuse of the concept of statistical significance remains one of the primary and most pervasive issues within the scientific community, particularly in the field of public health [1]. In general, there is a prevailing tendency to interpret the p-value as an objective measure capable of discerning scientifically significant results from those that are not [2]. However, as pointed out by its own creator, Ronald Fisher, and reiterated by numerous experts, including Sander Greenland, such a practice is entirely unfounded and contradicts the most recent evidence on the topic [3, 4]. When used appropriately, the p-value can assist in assessing the statistical relevance of findings in individual studies; however, its interpretation is subject to high and ineliminable margins of uncertainty. Furthermore, authors frequently blur the lines between the statistical surprise of a result and its clinical relevance, although these being two entirely separate aspects [5]. However, the purpose of this paper is not to reiterate concepts extensively discussed in the literature, but rather to conduct an investigation aimed at estimating recent trends in the adoption of these measures. Given that these kinds of errors can have severe consequences in the healthcare sector, including the approval of ineffective drugs or the rejection of effective ones, such an evaluation is both a priority and an urgent necessity. It should be clarified that the methodologies used to assess statistical significance in this paper adhere to the guidelines set forth by the American Statistical Association [6].

### Context and objectives

To ensure maximum transparency in data interpretation, the author briefly describes the motivations – and, consequently, the potential biases – that led him to undertake this research. The primary motivation is the author’s personal experience with these errors in the past, and his desire to contribute to preventing them in future studies. The secondary motivation arises from his roles as a peer reviewer and editor, where he has encountered this type of misuse in over 80% of the manuscripts analyzed. The research objective is to quantify the phenomenon in the current context as of October 2023. Specifically, this does not involve assessing the overall quality of the manuscripts, not even from a purely statistical perspective, but solely focuses on the use of Fisherian significance to inform public health decisions (also distinguishing between statistical significance and statistical effect size). It is also clarified that the aim is not to have a very precise estimate but rather a preliminary indication of prevalence. Furthermore, the author declares that he is not interested in the potential role of journals in the above mistakes (e.g., editorial requirements about P-values) but only in the overall infodemic scenario. Based on these considerations and the literature he has read on the subject, the author hypothesizes that a considerable number of misuses will be observed (potential bias).

## Methods

### Selection criteria and collection procedure

The PubMed database of the National Center for Biotechnology Information (NCBI) at the National Library of Medicine (NLM) was consulted for the study as it represents one of the most important repositories of scientific peer-reviewed medical articles in the world. To have a representative sample of the most recent trends, the current month was selected (from 11 September to 11 October, 2023). The search keyword was: “ANOVA” OR “regression” OR “t-test” OR “Chi square” OR “Mann-Whitney U test” OR “Kruskal-Wallis test” OR “Fisher’s exact test” OR “Logrank test” OR “Kolmogorov–Smirnov test” OR “Wilcoxon signed-rank test” OR “Dunnett’s test” OR “ANCOVA” OR “Levene’s test” OR “Friedman test” OR “Pearson correlation” OR “Spearman correlation” OR “Kendall correlation”. This choice was made considering the most commonly used statistical methods in the field of public health [7-10]. In addition, this increased the likelihood of finding studies containing analyses based on statistical significance. The search returned about 4,307 results. To get an approximate idea of a sufficient sample size, the formula for finite populations was used: n = (N·Z^2^·p·(1-p))/((E^2^·(N-1))+(Z^2^·p·(1-p))). Considering N = 4,307, Z = 1.96, p = 0.5, E = 0.1, n = 94 was obtained. In order to take into account possible exclusions, n = 110 was initially used. Through a random generator of integers from 1 to 4,307 with a uniform probability distribution, 97 studies with the following characteristics were selected: i) the study concerned public health topics, 2) the study contained quantitative results both in the text and the abstract, and 3) the study was peer reviewed. The general process is summarized in **Figure 1**.

**Figure 1.**
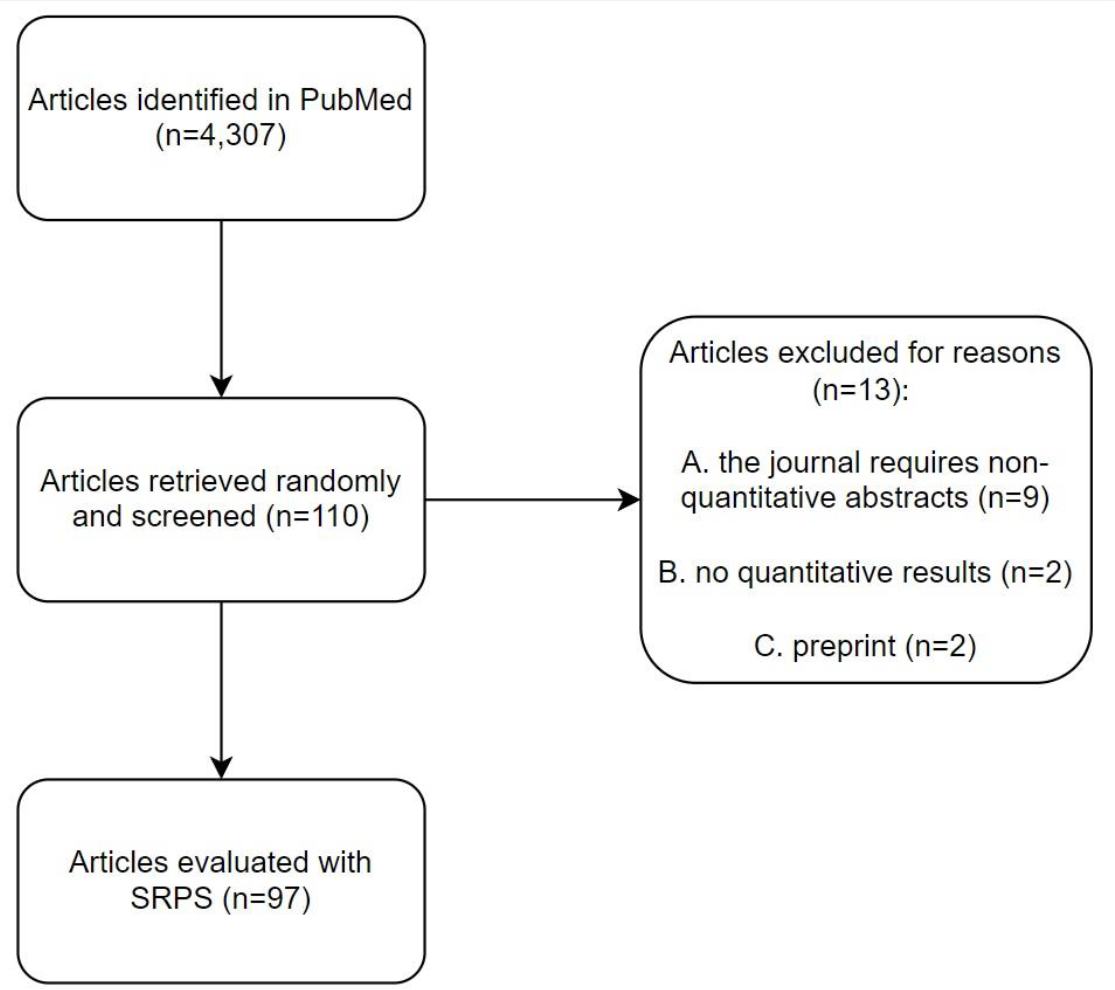
Articles selection and collection procedure flow chart.

### Evaluation scale

To evaluate the quality of the presentation of the results, four categories were defined according to the scheme shown below.

1. Significance continuity. 1 point is awarded if and only if statistical significance is measured continuously, i.e., the P-value is used as a continuous measure of the compatibility of the test result with the target hypothesis of the statistical model. In all other cases, i.e., when a threshold is adopted and/or when some results are referred to as “non-significant” and others as “significant,” 0 points are awarded.
2. Full p-values. 1 point is awarded if and only if all p-values are reported in full (unless P<.001) for all tests (regardless of the statistical significance of the results). 0.5 points are awarded when p-values are reported in full (unless P<.001) only for some measures (e.g., those considered significant); this includes mixed cases like P=.02 and P>0.05. 0 points are awarded when no p-values are reported in full. It is specified that notations like P<.05 fall into the latter situation.
3. Effect size measures. Confidence/credible intervals, standard errors, and specific measures like Cohen’s D or Hedges’ g have been included in this category. 1 point is awarded if and only if measures of statistical effect size have been reported for all conducted tests. 0.5 points have been awarded if measures of statistical effect size are reported for some of the tests conducted (regardless of the statistical significance of the results) or for individual measures before testing (e.g., reporting two means with respective standard errors but not a confidence interval of the difference after the z-test). 0 points are awarded when measures of statistical effect size are not reported for any test.
4. Best estimators. 1 point is awarded if and only if best estimators (e.g., correlation/regression coefficients, percentage differences, odd ratios, etc.) size are reported for all conducted tests (regardless of the statistical significance of the results). 0.5 points are awarded if best estimators are reported for some of the tests conducted (e.g., those considered significant). 0 points are awarded when measures of statistical effect size have not been reported for any test.
5. Standard assumptions (only for full texts). 1 point is awarded if and only if a complete assessment of the basic assumptions of the tests adopted is described and reported (in the paper or in supplementary materials). 0.5 points are awarded if a complete assessment of the basic assumptions is described but not reported. 0 points are awarded in all other cases.

This scale, called SRPS (statistical results presentation scale), was designed by considering the basic elements for a comprehensive evaluation of a statistical effect in a single study. These include the use of the Fisher approach (category 1), the reader’s ability to assess statistical significance independently (category 2), and the distinction between the size of the statistical effect and statistical significance (categories 3 and 4, as both descriptive statistics and standard errors are required). The maximum score obtainable was therefore 4 for abstracts and 5 for full texts. It is essential to specify that this paper does not investigate the methodological rigor with which the studies were conducted, but rather focuses solely on the basic statistical aspects (assuming that all other procedures were correct). The purpose is not to determine the overall methodological quality of the manuscript but rather the completeness and clarity in the application of basic statistical-inferential criteria. Indeed, these elements serve as the structural bedrock of the entire model, without which the overall research could potentially be undermined in terms of validity or interpretability. The reliability of the scale was tested by two independent raters on 50 papers. The obtained scores are reported in **Table 1**.

**Table 1.** Inter scale reliability of the SRPS tested on 50 manuscripts by 2 independent raters. The calculations were performed using RStudio, version 4.2.0, ‘psych’ library, ‘cohen.kappa’ function.

| Category | % agreement | Cohen's k | 95% CI | Weighted k | 95% CI | P-value |
| --- | --- | --- | --- | --- | --- | --- |
| 1 | 100 | 1.00 | 1.00 – 1.00 | 1.00 | 1.00 – 1.00 | <.0001 |
| 2 | 96 | 0.91 | 0.78 – 1.00 | 0.94 | 0.86 – 1.00 | <.0001 |
| 3 | 94 | 0.90 | 0.79 – 1.00 | 0.94 | 0.87 – 1.00 | <.0001 |
| 4 | 92 | 0.83 | 0.68 – 0.98 | 0.87 | 0.77 – 0.98 | <.0001 |
| 5 | 90 | 0.79 | 0.62 – 0.96 | 0.89 | 0.79 – 0.99 | <.0001 |

The SRPS was applied to both the abstract and the entire paper to highlight any differences. Indeed, while it is true that abstracts force authors to provide a partial representation of the results, it is also true that this constraint compels them to give more weight to the information they consider as most important; this can reveal any interpretative errors. In addition, the reading of an abstract could influence the interpretation of the entire manuscript (based on the confirmation bias mechanism) or, in more severe cases, even replace the reading of the manuscript. All data are reported in full at this URL: https://osf.io/3qgcs (DOI: 10.31219/osf.io/3qgcs).

### Statistical analysis

The paired Student t-test was employed to statistically assess the difference between the scores of the abstracts (a) and full manuscripts (f). The “assumptions” category was excluded to ensure a maximum score of 4 in both cases. Differences from the null value (f-a=0) and 0.5 (a-f=0.5) were investigated. Cohen’s D was applied to assess the statistical effect size. The assumption of data independence was considered satisfied due to the sampling method. The normality condition was deemed satisfied based on the central limit theorem [11]. The website “Statistics Kingdom” was used to perform the calculations [12].

## Results

### Abstracts analysis

None of the 97 studies adopted Fisher’s approach to describe the statistical significance of the results. The vast majority of studies did not report P-values in their entirety (52) or reported them only for results considered as statistically significant (38). The vast majority of studies did not report measures of effect size (48) or best estimators (17), or reported them only for results considered statistically significant (41 and 68, respectively). A complete summary is presented in **Table 1**.

**Table 1a.**
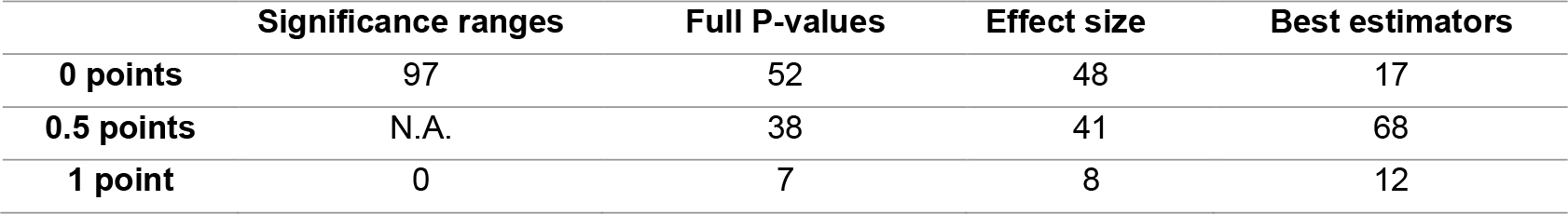
This table reports the scores achieved by the 97 abstracts included in the study. Legend: N.A. = not available.

### Full texts analysis

None of the 97 studies adopted Fisher’s approach to describe the statistical significance of the results. The majority of studies reported P-values in their entirety (69). Regarding effect size and best estimators, the range with the highest score had the most studies (43 and 64, respectively), although a substantial number of these did not report such measures (16 and 1, respectively) or only reported them for values considered as statistically significant (38 and 32, respectively). Finally, the vast majority of studies did not mention the basic statistical assumptions of the tests used (83) or merely explained how these were treated without reporting quantitative results or motivated arguments (9). A complete summary is presented in **Table 2**.

**Table 2.**
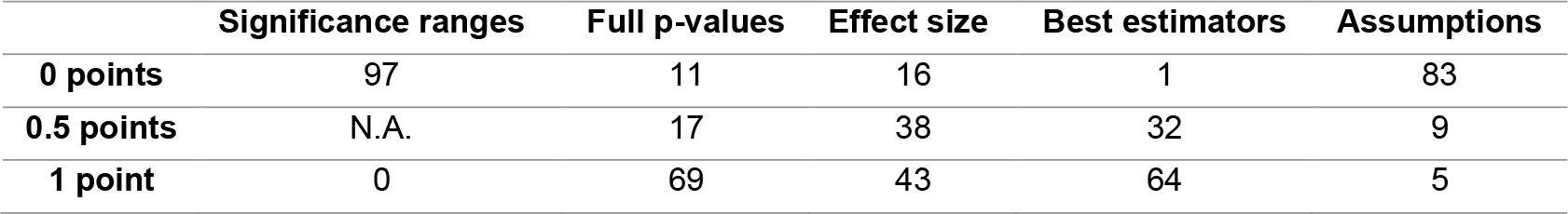
This table reports the scores achieved by the 97 full texts included in the study. Legend: N.A. = not available.

As shown in **Table 3**, the paired t-test revealed a highly significant and substantial difference between the scores obtained in the abstracts and the full papers.

**Table 3.**
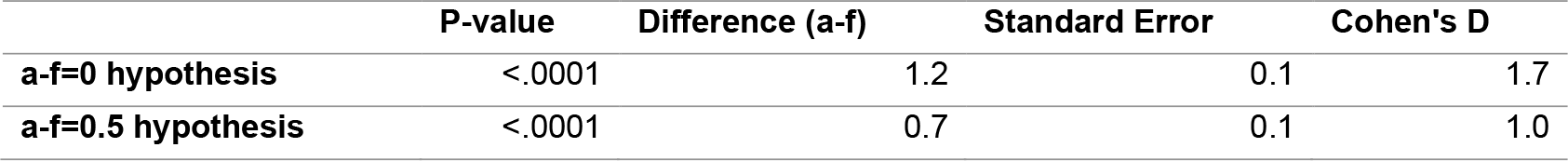
Results of the paired t-test comparing SRPS points between abstracts and full texts (df=96, priori power > 99%). Only the first four (from 1 to 4) categories were evaluated.

## Discussion

### Principal findings

This paper shows how the misuse of statistical significance is still highly prevalent in the field of public health. None of the 97 studies analyzed used Fisher’s approach to draw conclusions on the individual study at hand. At the same time, the dichotomous Neyman-Pearson approach, valid at most when employed to evaluate the total number of incorrect decisions in numerous well-conducted research, was mistakenly adopted to draw conclusions on the individual study [13]. However, in the majority of cases, albeit only in the full texts, all P-values, effect size measures, and best estimators were reported in their entirety, allowing the reader an independent evaluation. In this regard, the difference in result presentation between the full manuscripts and their abstracts is not solely explainable due to constraints imposed by the latter. On many occasions, even when dealing with few tests, only results with P<0.05 were presented and phrases indicating the statistical non-significance of other factors were reported without showing their respective P-values or effect size related measures. This behavior is in line with the concerning cognitive bias highlighted by the statistical community [14, 15]. In addition to this, almost all the studies provided no framework for assessing the validity of the statistical assumptions underlying the tests employed, making the reported findings extremely susceptible to compromising margins of interpretative errors [1]. This evidence highlights the little importance generally given to the fundamental hypotheses of the model used, which are actually as essential as the target hypothesis and deserve the same degree of analytical attention [2].

### Practical implications

While governmental and health agencies such as the World Health Organization, Centers for Disease Control and Prevention, Food and Drug Administration, and European Medicines Agency have their internal evaluation committees dedicated to ensuring the clinical efficacy of treatments and drugs, these widespread errors and uncertainties in the field of clinical research can not only propagate a marked infodemic – as often witnessed during the COVID-19 pandemic – but also result in a wastage of resources, such as the prolonged funding for studies with exaggerated outcomes [16, 17]. In fact, sensationalistic expressions are aimed at increasing the perception of the study’s relevance beyond its actual findings, i.e., to boost the number of citations and success, a crucial factor in securing research funding and even institutional roles [18]. For instance, the scientific community has been decrying the widespread practice of p-hacking for decades, although the consequences of this misconduct are a subject of debate [19, 20]. Given that, as highlighted by the undersigned and various experts in the field as well as supported by these findings, there is a furious resistance to changing these scientifically unsound practices, the author of this manuscript calls for academic journals to begin mandating scientific standards that align with the latest statistical evidence advocated by organizations such as the American Statistical Association. Furthermore, journal editorial policies should assign equal weight to both positive and negative findings. This must be done in the name of scientific and medical ethics since it is an essential step toward conducting unbiased investigations. Based on this, the following basic recommendations are proposed. First, only if all test assumptions are sufficiently met (a methodological aspect to be extensively discussed in the manuscript, especially when dealing with clinical results), academic journals should explicitly and compulsorily require that P-values be treated as a continuous measure of the compatibility between the test result and the target hypothesis (e.g., null hypothesis) when dealing with single-study-based conclusions. Specifically, P-values close to 1 indicate high compatibility, while P-values close to 0 indicate low compatibility. Second, academic journals should explicitly and compulsorily require that the effect size be treated as a completely separate aspect from statistical significance. Third, academic journals should explicitly and compulsorily require that authors refrain from using sensationalistic expressions when presenting results, especially if the latter stem from statistical analyses. As a matter of fact, statistics can provide – when hypotheses are well-targeted, i.e., motivated by evidence of other kinds – further evidence in favor of or against a phenomenon but can never, in any way, prove or disprove its existence. In this regard, it must be emphasized that the P-value refers only to the test result and not the phenomenon under investigation in itself. Finally, academic journals should explicitly and compulsorily require that public health recommendations must be provided only after an analysis of data sensitivity, biases, confounding factors, risks, costs, and benefits, and not on the P-value or any other statistical indicator [21-23]. In this regard, guidelines and checklists previously discussed in the literature (e.g., SAMBR) can be helpful [24-26]. Moreover, it’s also worth noting that there are international initiatives aimed at improving the quality and transparency of health research, such as EQUATOR [27].

### Proposal

Based on the above, the following guidelines are proposed for reporting basic statistical analyses comprehensively:

1. In the manuscript, provide a brief explanation of how the tests were performed and how the related assumptions were examined, specifying – if necessary – that methodological details are documented in a supplementary file. Authors opinions and expectations should be clearly stated in order to openly acknowledge potential biases.
2. Present all calculations and procedures used to validate the adopted tests (including their assumptions). Quantitative and qualitative results, including graphs, should be fully reported so that readers can independently and easily assess their reliability. Indeed, a statistical test is reliable if and only if the underlying assumptions are true (or sufficiently met). This point can be addressed directly in the manuscript or, if necessary, in a supplementary file.
3. Present, at least in the full text, all P-values and effect size measures, regardless of significance or other properties.
4. Avoid the dichotomous use of the terms “significant” and “non-significant” since they are unfounded when drawing conclusions about an individual study. Instead, use the P-value as a continuous measure of the compatibility of tests results with the target hypotheses (after validating their assumptions).
5. Additionally, though not addressed in this study, it is crucial to consider the implementation of more advanced techniques (e.g., adjustment for multiple comparisons and sensitivity analysis) depending on the research purpose and scenario [28]. This includes the adoption of multiple hypotheses (e.g., difference = 0, difference = 0.5, etc.) or multiple confidence intervals (e.g., notation 90/95/99%-CI) [2]. The references mentioned above can be useful for this purpose.

### Limitations and their potential impact

This study has some limitations that should be taken into account. Firstly, the sample was collected over a very recent but limited period. It is possible that there are trends or periodic oscillations to consider, even though the author is not aware of them and does not find valid reasons to suspect their existence. Secondly, the study focused on the most commonly used statistical methods in public health but did not consider other statistical methods that might be adopted in this field. However, since i) the goal is to provide a simple overview and ii) the primary statistical methods have been included, the author believes that this potential limitation does not have a practical impact. Thirdly, part of the study relied on a newly developed evaluation scale (SRPS), which, while tested for reliability, has not been extensively validated. Nevertheless, the interpretation of this scale is very straightforward and, in any case, allows for easy independent reading by the reader. Fourthly, it is possible that some researchers may not have used acronyms (e.g., ANOVA) but instead provided an extended description (e.g., analysis of variance). However, the author is not aware of any behavioral distinctions between those who use acronyms and those who do not. For this reason, he considers the sample to be sufficiently representative.

## Conclusion

These findings provide preliminary evidence of widespread and severe shortcomings in the use and interpretation of statistical significance measures in clinical and public health research during 2023. This is consistent with decades of criticism from epidemiologists and statisticians, including respected international organizations. Such errors can result in highly misleading interpretations, posing a threat to public safety. As a result, it is essential for academic journals to demand higher scientific quality standards.

## Data Availability

All data produced in the present work are contained in the manuscript or this supplementary file: https://osf.io/3qgcs

https://osf.io/3qgcs

## Ethical Considerations

The author declares that he has no conflicts of interest.

